# LncRNA PVT1 in brain injury induced by deep hypothermia and low flow

**DOI:** 10.1101/2024.08.13.24311905

**Authors:** Yuzhong Yang, Siyu Ma, Xiaodong Zang, Xuming Mo

**Affiliations:** Department of Cardiothoracic Surgery, Children’s Hospital of Nanjing Medical University, Nanjing, China; The First Affiliated Hospital of USTC, Division of Life Sciences and Medicine, University of Science and Technology of China, Hefei, Anhui, China

**Keywords:** deep hypothermic low flow, ischemia-reperfusion injury, long non-coding RNA PVT1, miR-148a-3p, MKL1

## Abstract

**Aims:** To analyze the role of lncRNA PVT1 in cerebral ischemia-reperfusion (I/R) injury induced by deep hypothermia low flow (DHLF).

**Methods and results:** A total of 72 lncRNAs were differentially expressed in the brain tissue of DHLF mice. PVT1 expression was significantly downregulated in DHLF mouse brain tissue, preoperative and postoperative blood samples from children undergoing DHLF extracorporeal circulation, and hOGD-treated cells. In the mouse model, the DHLF group with PVT1 overexpression had heavier brain tissue damage than the control group; apoptosis rate, reactive oxygen species level and caspase-3 enzyme activity were significantly higher in the lenti-PVT1 group than in the lenti-control group. Compared those in the lenti-control group, the total distance traveled, distance of action in the center, number of entering the center, average speed of walking reduced, and the distance of walking in the periphery and and peripheral walking distance increased in the lenti-PVT1 group. The dual luciferase reporter gene assay verified the possible binding sites between PVT1, miR-148a-3p and MKL1. In the animal model, cellular model, and blood samples of children experiencing DHLF, miR-148a-3p expression increased and MKL1 expression decreased. In experimental studies in vivo and in vitro, PVT1 and MKL1 expression increased, and miR-148a-3p expression decreased. Meanwhile MKL1 inhibitor CCG1423 reversed the apoptosis in neuronal cells.

**Conclusion:** PVT1 may adsorb miR-148a-3p to regulate the expression of MKL1, a downstream gene of miR-148a-3p, a mechanism promoting the apoptosis of neuronal cells in DHLF mice.

## 1. Introduction

Ischemia reperfusion injury (IRI) often emerges as a complication in patients with congenital heart disease (CHD) receiving surgery under extracorporeal circulation. About 220,000 children with CHD are annually reported in China [1]. Deep hypothermic low flow (DHLF) technique has been invented to mitigate IRI caused by extracorporeal circulation [2]. In patients with complex congenital heart disease (CCHD), the surgery has a long duration, and the organs experience a long period of ischemia and hypoxia. Although protected by the DHLF technique, I/R still damages the central nervous system (CNS), especially the brain [3,4]. Signs of CNS damage include seizures, psychomotor disorders and cognitive impairment [5,6]. Therefore, the genes and signaling pathways involved in cerebral IRI after DHLF remain to be clarified to design related treatment options.

Studies have shown that impairment of mitochondrial function, dysregulation of apoptotic and autophagic pathways, and abnormalities in signal transduction are involved in IRI [7]. However, the specific mechanisms of tissue damage caused by I/R have not been fully elucidated. Recent studies have shown that non-coding RNAs (ncRNAs) play an regulatory role in the IRI, including long non-coding RNAs (lncRNAs), microRNAs (miRNAs), and circular RNAs (circRNAs) [11]. lncRNAs can regulate gene expression at the epigenetic, transcriptional, and post-transcriptional levels in various pathological processes, such as autophagy, necrosis and apoptosis [12]. In recent years, lncRNAs and IRI have received much research attention. lncRNA ROR represses miR-135a-3p to increase the expression of rock1/2, a mechanism that may induce apoptosis [13]. Upon ischemia, lncRNA CHRF is upregulated and miR-126 downregulated, and silencing lncRNA CHRF can inhibit ischemic brain injury and ameliorate neurological deficits in animal models [14]. Zhang et al. showed lncRNA ZFAS1 overexpression suppressed I/R-induced inflammation, oxidative stress and apoptosis in neuronal cells, thus ameliorating neurological dysfunction and neuronal injury in an MCAO rat model [15].

A close link lies between neuronal apoptosis and brain injury after I/R. In the brain tissue, ischemia and hypoxia attenuate cellular energy metabolism, accompanied by damage to subcellular organelles. Therefore, finding effective molecular targets to inhibit neuronal apoptosis is the key to the treatment and prevention of IRI in brain tissue. In this study, we proposed to find the key molecules regulating apoptosis and to investigate its specific molecular mechanisms.

## 2. Materials and methods

### 2.1. Experimental materials

#### 2.1.1 Experimental animals

Male C57BL/6 mice (3-4 weeks old, 8-10 g) were sourced from the Experimental Animal Center of Nanjing Medical University, raised in an SPF-grade environment with constant temperature and humidity in a 12-hour light/12-hour dark cycle. The experimental procedures were conducted in strict accordance with the guidelines in the Guide for Ethical Review of Laboratory Animal Welfare. The experimental operation process strictly abided by the regulations of the Experimental Animal Management Committee of Nanjing Medical University.

#### 2.1.2 Experimental cells

Neuro-2a cells (N2a, mouse neuroblastoma cells), HEK293T cells (human embryonic kidney cells 293) were purchased from the Chinese Academy of Sciences (Shanghai Institute of Biological Sciences Cell Resource Center).

#### 2.1.3 Experimental reagents

Annexin V-PE Apoptosis Detection Kit, Reactive Oxygen Species Detection Assay Kit, one-step TUNEL Apoptosis Assay Kit, Luciferase Reporter Assay Kit (Biyuntian Biotechnology Co., Ltd.); PVT1 overexpression lentivirus, miR-148a-3p agomir and miR-148a 3p antagomir (General Biosystems Ltd.); rabbit-derived anti-β-actin, rabbit-derived anti-MKL1 antibody (ABchlonal Technology), etc.

#### 2.1.4 Primer sequences

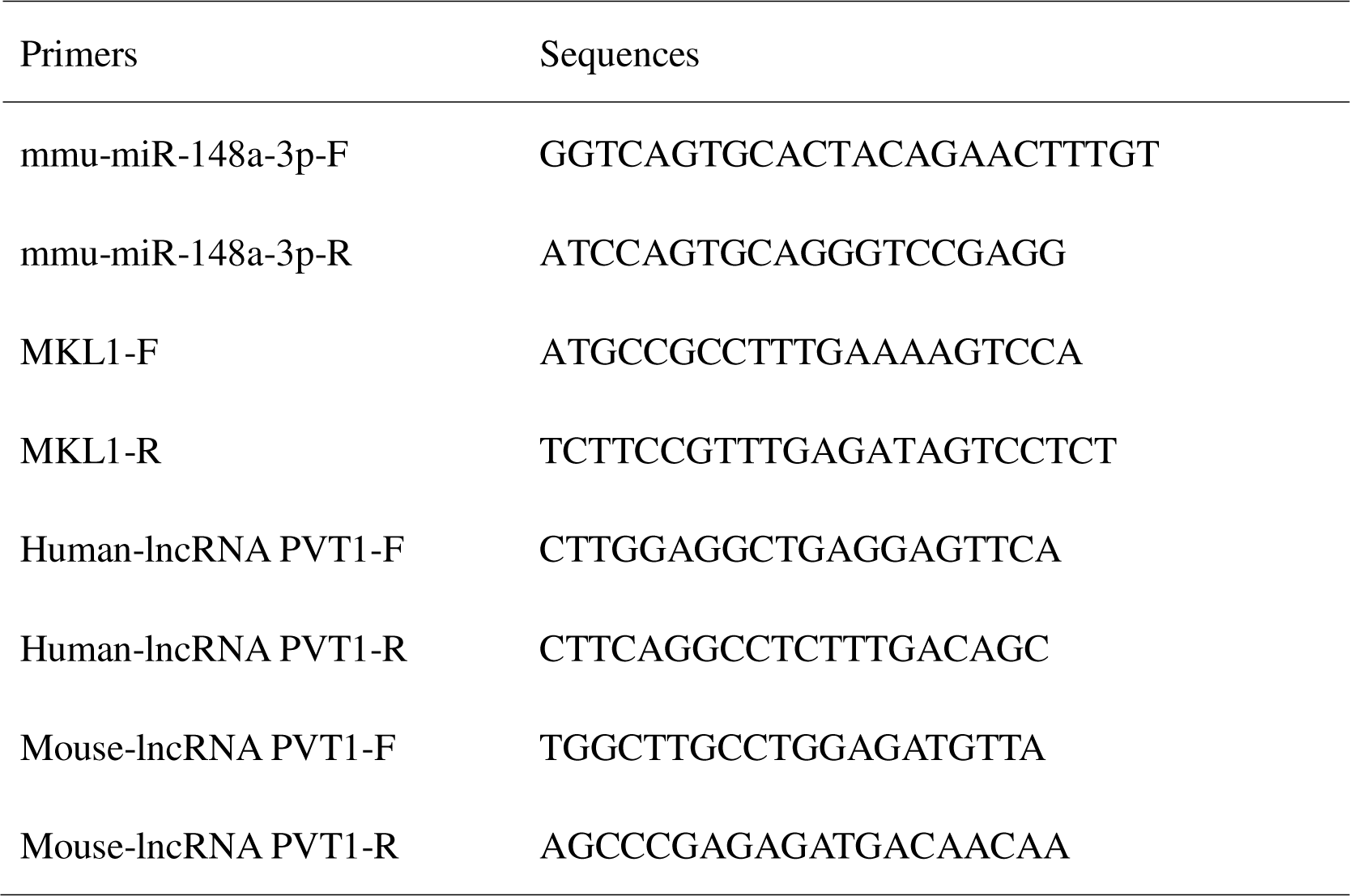

### 2.2. Experimental methods

#### 2.2.1 Collection of blood samples

Informed consent was obtained from the child’s guardians or legal delegates. Blood was sampled before and within 24 h after surgery from September 2019 to December 2021.Informed consent was obtained from the legal guardian of the children, and the protocol was approved by the Institutional Ethics Committee of Children’s Hospital of Nanjing Medical University.

Inclusion criteria

1. Preoperative diagnosis of CCHD by imaging.
2. CCHD treated with DHLF.

Exclusion criteria

Preoperative abnormalities, including abnormal neurological development, hematologic disorders, chromosomal abnormalities, and severe lesions of other organs, etc.

#### 2.2.2 Establishment of a DHLF mouse model [2,3,16]

C57BL/6 male mice (3-4 w) were intraperitoneally injected with 5% chloral hydrate (0.35-0.5 mg/kg). The mouse was fixed in a supine position on the operating table. Its neck was disinfected, and an incision was made in the median neck. Bilateral common carotid arteries and nerves and other tissues were separated. The mouse was placed on an ice bed, and the rectal temperature was measured by platinum resistance electronic temperature sensor. When the temperature dropped to 18□-19□, the bilateral common carotid arteries were clamped with non-invasive fiber artery clips for 120 min. The rectal temperature of the mouse was maintained at 18□-19□ throughout the procedure. Afterward, the bilateral common carotid arteries were opened, the neck incision was disinfected, and the incision was sutured. The mouse was placed in a neonatal incubator to rewarm. The body temperature was allowed to recover with a rate of ≤0.5□/min to 32□ within 30 min, after which the constant temperature of the warming box was maintained and the temperature was slowly increased until recovery. In the sham group, the operation was performed in the same manner except that the bilateral common carotid arteries were not blocked and the temperature was not lowered.

#### 2.2.3 Monitoring of regional cerebral blood flow (rCBF) in mice [17,18]

C57BL/6 male mice (3-4 w) were randomly divided into DHLF (n=6) and sham groups (n=6). 5% chloral hydrate was injected intraperitoneally (0.35-0.5 mg/kg). The mouse was fixed in a prone position on the operating table. Parietal skin was prepared. Drill a hole with a diameter of 1 mm at 1 mm behind the skull anterior halogen and 2 mm to the left / right of the midline. A laser Doppler microfiber probe (0.5 mm in diameter) was placed in contact with the cerebral cortex through the above hole and fixed to the skull. The cerebral blood flow was monitored using Perisoft Version 2.50 Perimed software and recorded before and at 18±0.5°C, 0-5 min, 25-30 min, 55-60 min, 115-120 min, and 0-5 min after extracorporeal circulation at 18±0.5°C.

#### 2.2.4 Establishment of a PVT1 overexpression mouse model [19]

The mice were randomly divided into 3 groups (sham group, lenti-PVT1 group: lentivirus overexpression group and lenti-control group: overexpression empty vector group). Chloral hydrate 5% was injected intraperitoneally (0.35-0.5 mg/kg). The mouse was placed in a prone position and fixed on an operating table. After parietal skin disinfection, an incision 5 mm in length was made along the sagittal plane at the midpoint of the posterior margin of both ears. The skull was punctured with a needle of 1 ml syringe to make a hole about 1 mm in diameter, located 1-2 mm outside the left or right corner of the fontanel and 1-2 mm backward (turn the needle slowly at the fixed point). The needle was inserted into the ipsilateral medial canthus at a depth of about 2-3 mm (fix the mouse head and microliter microsampler). The injection rate was 2 μl/10 min; after the injection, the needle was maintained for 10 min, then pulled out. The injection hole was pressed and the wound was sutured.

#### 2.2.5 Deep hypothermic oxygen glucose deprivation (hOGD) cell model

Cells were cultured to a confluence of 70%-75%. Serum-free medium was substituted with complete medium and placed in an incubator containing 5% CO2 and 95% air at 37°C for 2 h. The medium was gently aspirated with a pipette and 2 ml of PBS was added to each dish, which was repeated for three times. Then, 2 ml of fresh DMEM sugar-free medium was added. The cells were incubated in a triple-air incubator with 95% N2, 5% CO2 and 1% O2 for 24 h. The medium was gently aspirated with a pipette and 2 ml of PBS was added to each dish, which was repeated three times. Next, 2 ml of DMEM complete medium was added to each well, and the cells were incubated in an incubator with 5% CO2 and 95% air at 37°C for 24 h.

#### 2.2.6 Lentiviral plasmid transfection in vitro

The transfection was performed strictly according to the protocol in the kit.

#### 2.2.7 Total RNA extraction, RNA reverse transcription, real-time fluorescence quantitative PCR

Total RNA extraction, reverse transcription, and real-time fluorescence quantitative PCR were performed according to manufacturer’s instructions. Reverse transcription products were used immediately for qPCR or stored at -20°C within six months. For usage in a longer, the products were stored at -70°C after splitting. cDNA should avoid repeated freeze-thawing.

#### 2.2.8 TTC staining

TTC staining was performed according to manufacturer’s instructions. Image J software was used to measure the volume of non-infarcted and infarcted parts. The volume= average of the front and back areas×section thickness (2 mm), and the volumes of consecutive sections were summed as the total volume.

#### 2.2.9 Neurological deficit scoring

The mice were assessed for neurological function at 1 week after the establishment of the mouse DHLF model. A modified neurological deficit scale (total 14 points) was used:

1. Tail-Hanging Test (THT) The tail of the mouse was gently left above the ground by 10 cm. Each of forelimb flexion, hindlimb flexion or head rotation deviated from the central axis by more than 10 degrees was scored by 1 point.
2. Flat movement test The mouse was placed on a flat floor. Its movement scored 0 (walking normally); 1 (cannot walking in a straight line), 2 (walking in a circle), 3 (stumbling during walking).
3. Sensory test The auricle and cornea were stimulated. If the corneal reflex or auricular reflex disappeared, 1 point was given.
4. Balance beam test The mouse was gently placed on the balance beam and allowed to walk freely. Its performance scored 0 (normal walking), 1 (holding on to the balance beam), 2 (one foot falling down), 3 (two feet falling down or rotating in a circle), 4 (being able to maintain balance for more than 40 s), 5 (being able to maintain balance for more than 20 s), 6 (difficult to maintain balance and fall within 20 s).
5. Total score=the sum of the four scores.
6. Degree of neurological injury

Mild injury: 1-4 points; moderate injury: 5-9 points; severe injury: 10-14 points.

#### 2.2.10 Nuclear magnetic resonance imaging

The mouse model of DHLF was established. Every 6 mice were randomly taken as a group, anesthetized, prone-positioned, head-fixed. Cranial MIR was performed with a layer thickness of 1 mm and a layer spacing of 1 mm, acquisition matrix of 256×256, FOV of 20×20 mm, TR of 5000 ms, TE of 16.25 ms, and 4 times of excitation. Image J software was used for image processing and analysis, and cerebral infarction volume was calculated.

#### 2.2.11 Open field test

The DHLF mouse model was established. At day 28 or 29, every 12 mice were randomly taken as a group for open field test. The steps were as follows.

Parameters of the field box: 25 cm height, 50×40 cm bottom, black inner wall. A camera was placed 1 m above the box to ensure that its field of view can observe the internal area of the open field. The mouse was placed in the open field box, and the experimental personnel and equipment (such as computers) were placed in another room to reduce interference with animals (e.g., laboratory background noise below 65 dB).

#### 2.2.12 Luciferase reporter gene assay

HEK293T cells were used in the assay according to related instructions. The fluorescence value (Luc fluorescence value) was measured; the ratio of Rluc fluorescence value to Luc fluorescence value in each well was calculated. The ratio was compared with that in the control wells.

#### 2.2.13 Reactive oxygen assay and flow cytometric assay

The assay was performed according to kit instructions.

#### 2.2.14 Total protein extraction, quantification, and protein blotting

The concentration of lower layer gel was set according to the protein molecular weight size. Electrophoresis conditions (15 mA for concentrated gel, 20 mA for separation gel, or 100-120 V at constant pressure). Transfer membrane conditions: 200 mA for 1-2 h, or 100 V for 1-2 h under a constant pressure.

#### 2.2.15 Immunofluorescence assay

The assay was performed strictly according to the instructions. Immunofluorescnence was observed under a laser confocal microscope and analyzed by Image J software.

#### 2.2.16 Statistical analysis

The experimental data were expressed as mean ± standard deviation, and SPSS and Graphpad Prism 8 software were applied for statistical analysis. One-way ANOVA was performed for comparison between groups, and t-test for two-by-two comparison. p<0 .05 was considered a statistically significant difference.

## 3. Results

### 3.1. Local cerebral blood flow in the DHLF mouse model

In the mouse model of DHLF, rCBF was monitored at 7 time points: before, 18-19□, 0-5 min, 25-30 min, 55-60 min, 115-120 min during and 0-5 min after extracorporeal circulation (Table 1). Clinically, when the rectal temperature is 18-20°C, the perfusion flow rate in extracorporeal circulation should be reduced from 120 ml/kg/min to 20-25 ml/kg/min, letting the blood supply flow to the brain account for about 20% of the normal state. Therefore, to simulate the changes in cerebral blood flow during the surgery under extracorporeal circulation, the bilateral common carotid arteries were blocked to establish a low perfusion of brain tissue. The cerebral blood flow in the mouse during DHLF decreased to 15-20% of the normal, similar to that in children receiving DHLF (Table 1). H&E staining showed that the cells in the sham group were densely arranged with obvious nuclei and without abnormal neurons (Figure 1, a&b). In the DHLF group, the neurons were sparse and edematous, with poorly defined nucleoplasm, inconspicuous nucleoli, and solid, dark-stained nuclei. Nisei staining showed that the cells in the sham group were densely packed with abundant Nisei vesicles, with obvious axons, and the cytoplasm was stained dark blue (Figure 1a, b). In the DHLF group, the neurons were sparse and edematous, with distorted and swollen cells, fewer Nisei vesicles, and inconspicuous axons. Apparent neuronal cell damage was observed. Therefore, the cerebral blood flow in the DHLF model could simulate that in children with DHLF.

**Figure 1.**
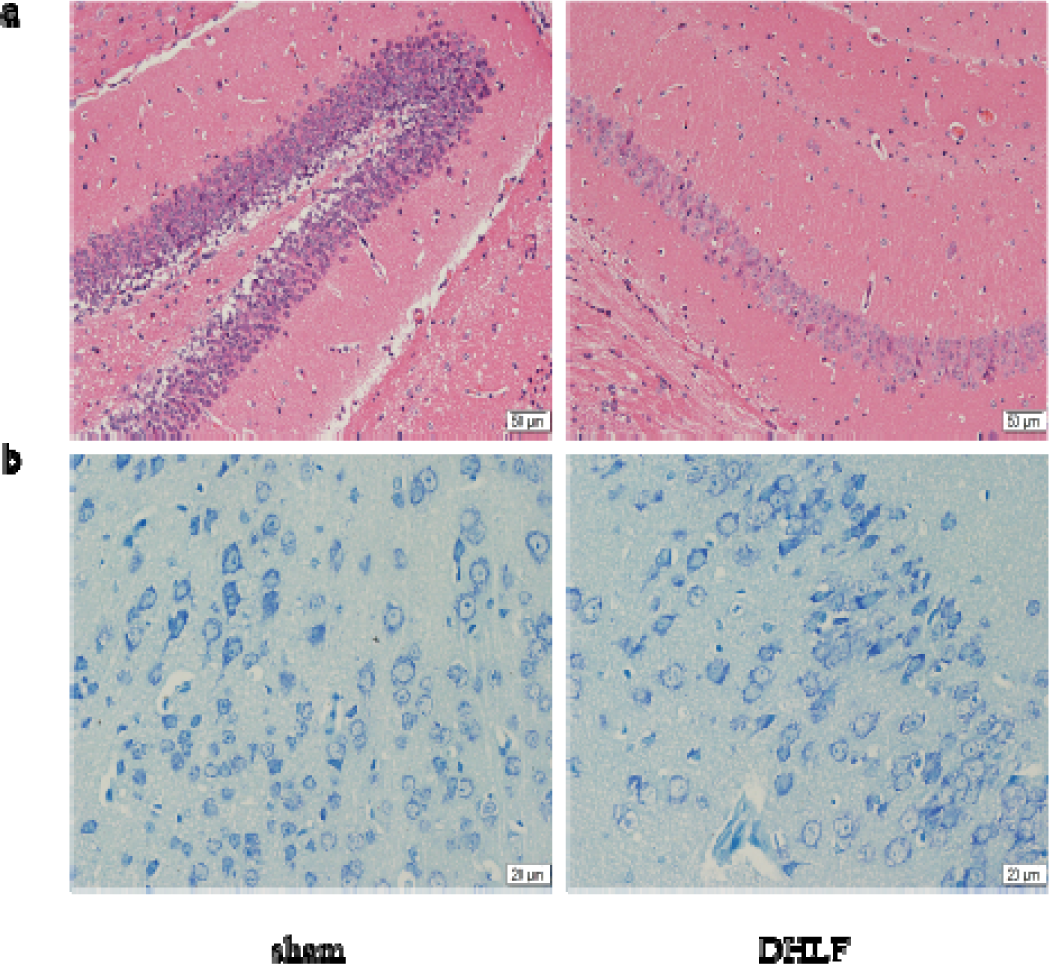
HE staining and Nissl staining of brain tissue in the DHLF mouse model. a: HE staining; b: Nissl staining. (DHLF: deep hypothermic low flow model group).

**Table 1.** Changes in local cerebral blood flow of mice in DHLF model (%)

| group | No<br>cooling | 18±0.5□ | time |  |  |  | After<br>ischemia-r<br>eperfusion<br>0-5 min |
| --- | --- | --- | --- | --- | --- | --- | --- |
|  |  |  | 0-5 min | 25-30 min | 55-60 min | 115-120min |  |
| sham | 97.5±1.2 | 97.6±1.3 | 97.1±1.4 | 97.8±2.1 | 98.2±1.2 | 97.5±1.2 | 97.6±0.9 |
| DHLF | 97.9±0.6 | 45.8±1.2* | 21.5±2.3* | 19.2±2.6* | 17.8±3.2* | 17.5±2.0* | 42.1±2.0* |

\* p <0.005 vs sham , decreased cerebral blood flow , n= 6。

### 3.2. RNA-seq data screening and qPCR validation

To analyze the changes lncRNA expression in brain tissue after DHLF surgery, high-throughput sequencing was performed in the sham and DHLF groups (n=5). 97924839 and 95944704 raw sequences were obtained and filtered in the sham and DHLF groups, respectively. A total of 144,042 transcripts were obtained. Then, 405 annotated and unannotated lncRNAs were obtained by filtering the transcripts through exon number, coding potential, length and expression level (Figure 2a, b). Between the two groups, 72 lncRNAs were differentially expressed, including 38 lncRNAs up-regulated and 34 down-regulated (Figure 2c). PVT1 expression was significantly altered in the DHLF group. The expression profiles obtained by RNA-seq were verified using real-time PCR technique. As shown in Figure 2d, the most highly up or down-regulated lncRNAs in the DHLF mouse model were consistent with the sequencing results.

**Figure 2.**
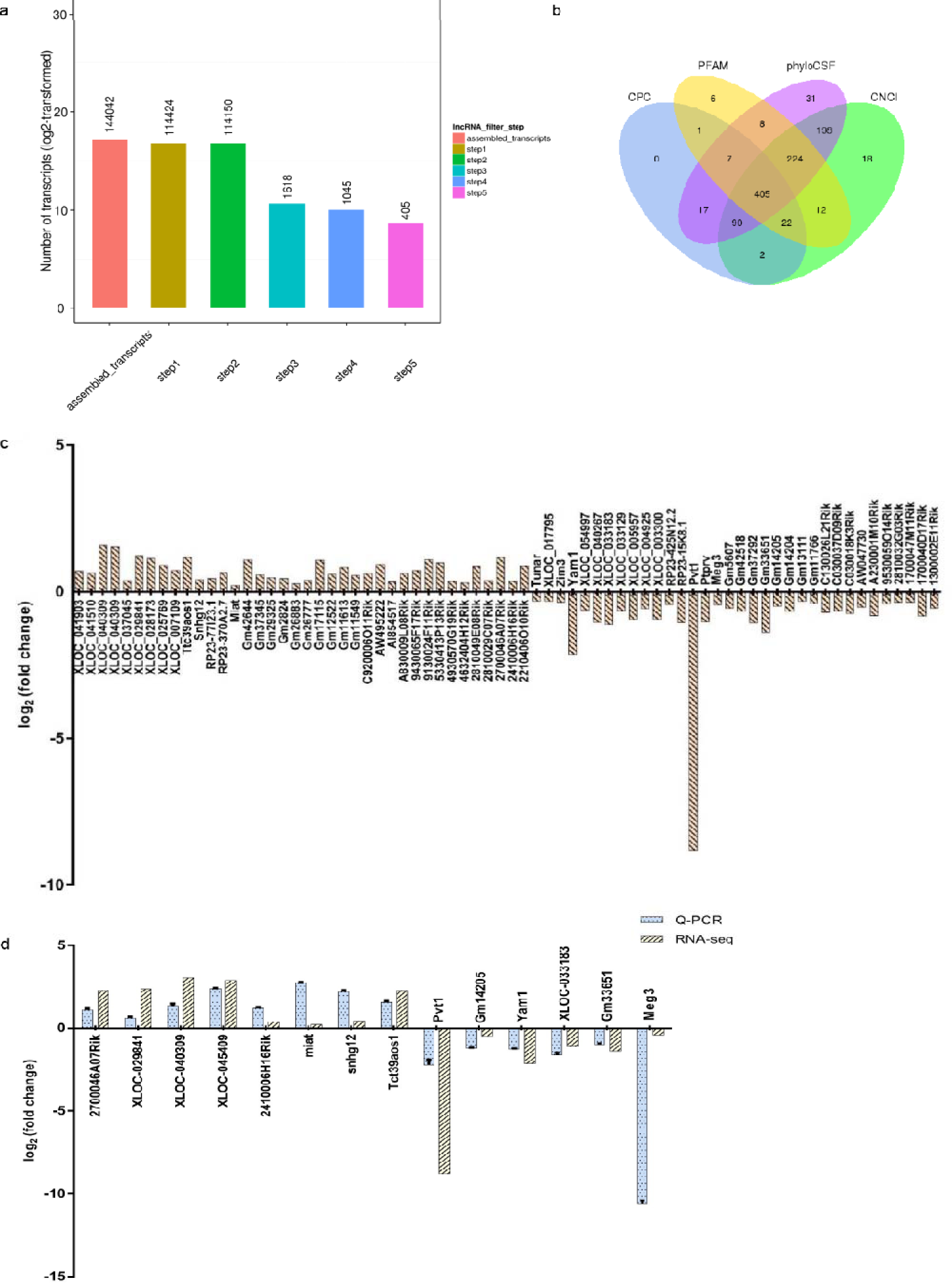
Differentially expressed lncRNAs in brain tissue of deep hypothermic low-flow mouse model. a: number of transcripts; b: results of four coding potential screening software; c: 72 differentially expressed lncRNAs (X-axis: different transcripts; Y-axis: logarithm); d: validation of the changes of differentially expressed lncRNAs by real-time PCR (n=20).

### 3.3. PVT1 interaction network

The PVT1 interaction network was generated with Cytoscape software based on the results of RNA-seq expression profiles (Figure 3). arpp21, Pde7b, Sh3rf2, Gad1, Nfib, Tbc1d1d, Enox2, Cdhr4 and 0610040J01Rik functioned as major subcenters of PVT1.

**Figure 3.**
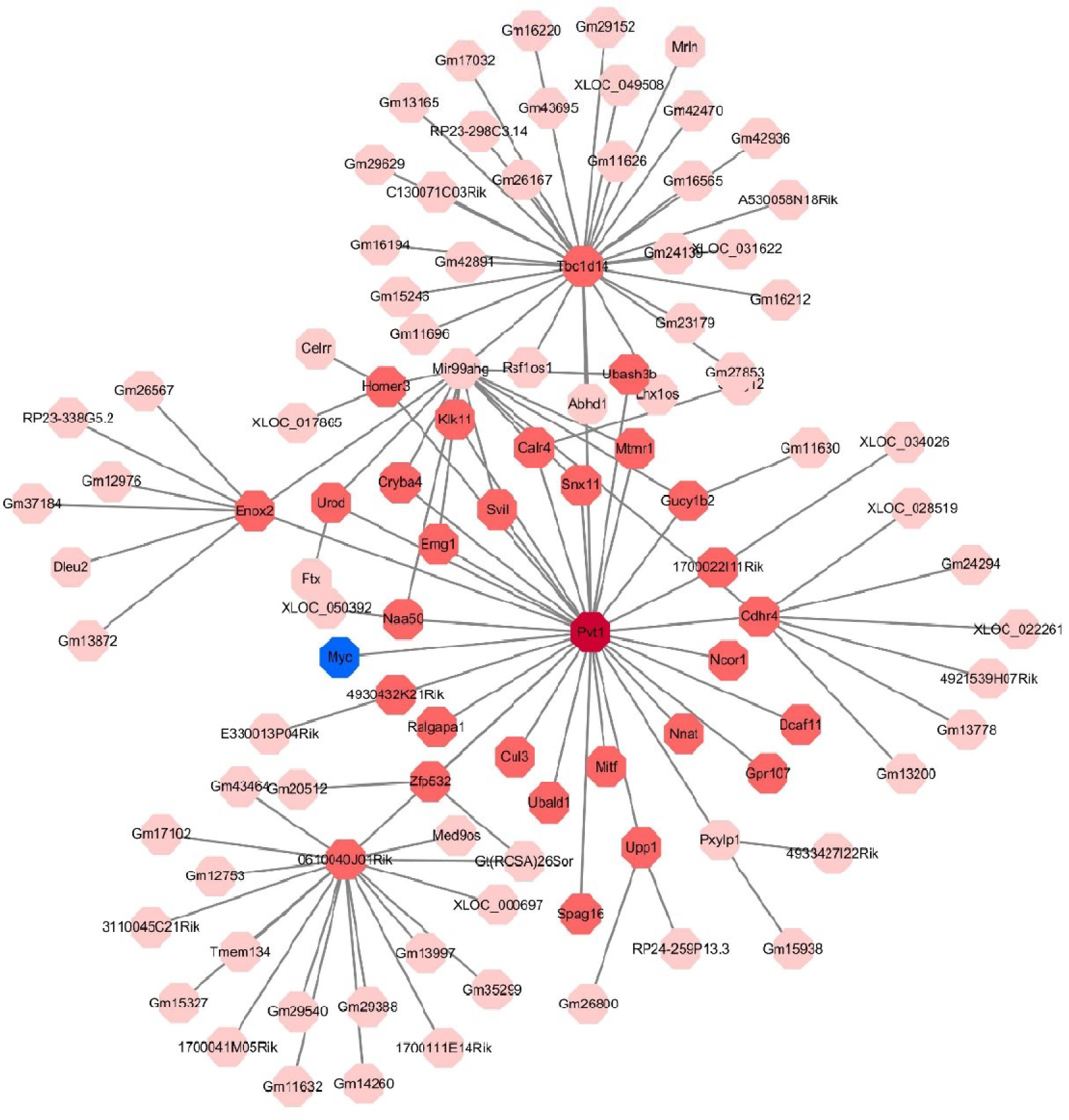
gene interaction network of lncRNA PVT1

### 3.4. Serum PVT1 expression in children with CHD

Pre and post-operative blood samples were collected from children undergoing DHLF, and changes in PVT1 expression were verified using qPCR. Table 2 shows the clinical and demographic characteristics of all children included in this study. As shown in Figure 4, the expression level of PVT1 in the blood was significantly decreased after DHLF, compared with that before DHLF. This change was consistent with that in PVT1 expression in the DHLF mouse model.

**Figure 4.**
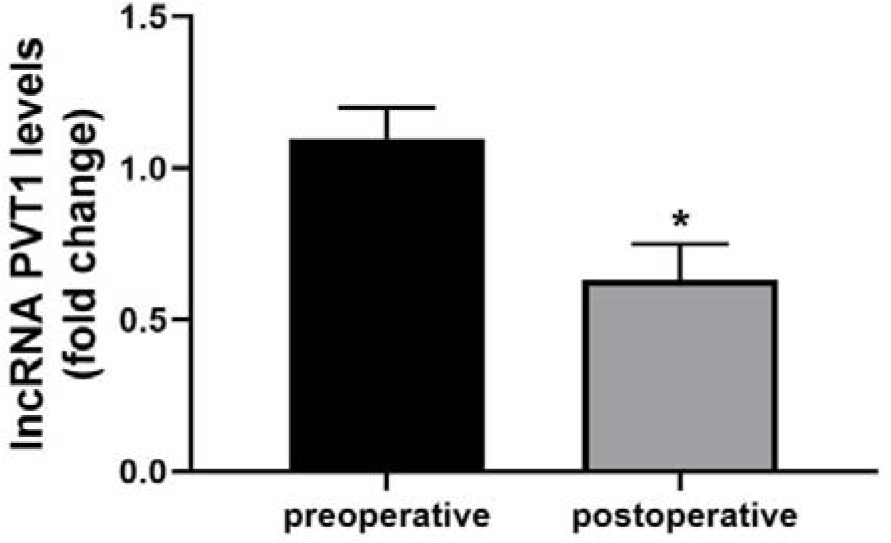
Expression of lncRNA PVT1 in the blood of children with congenital heart disease. (*p<0.05)

**Table 2.** Table of clinical and demographic characteristics of the children undergone DHLF.

| category |  |
| --- | --- |
| age ( day ) | 30.00 ( 12.00-75.00 ) |
| gender |  |
| male | 16 ( 53.33% ) |
| female | 14 ( 46.67% ) |
| weight ( Kg ) | 3.15 ( 2.50-5.25 ) |
| extracorporeal circulation time ( min ) | 120.55 ( 70.20-210.50 ) |
| aortic occlusion time ( min ) | 50.20 ( 30.17-90.25 ) |
| Body temperature ( °C ) | 20.20 ( 18-22 ) |
| ICU treatment time ( day ) | 12.3 ( 7.00-35.00 ) |
| Total length of stay ( day ) | 48.20 ( 45.20-72.30 ) |
| classification of congenital heart disease( case ) |  |
| coarctation of aorta | 8 |
| interrupted aortic arch | 4 |
| total anomalous pulmonary | 10 |
| venous drainage |  |
| double outlet of right ventricle | 6 |
| complete transposition of great arteries | 2 |

### 3.5. Expression of PVT1 in deep hypothermic glucose oxygen deprivation cell model

A deep hypothermic glucose oxygen deprivation-reoxygenation (hOGD-R) cell model was established. The changes in the expression of PVT1 were verified using qPCR. As shown in Figure 5, the expression level of PVT1 was significantly decreased in the hOGD-R group than in the control group. This result was consistent with that in the DHLF mouse model and blood samples of children experiencing DHLF.

**Figure 5.**
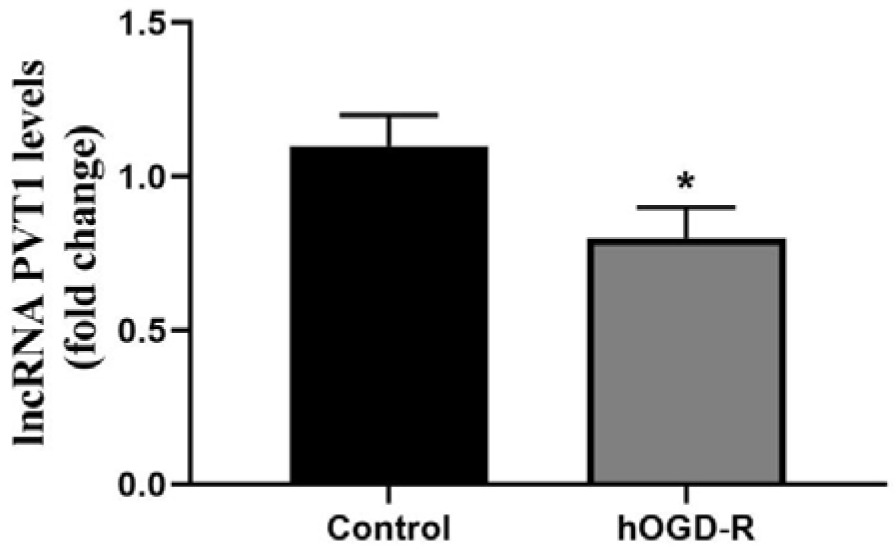
Expression of lncRNA PVT1 in the deep hypothermic glucose oxygen deprivation cell model. (*p<0.05)

### 3.6. Effect of PVT1 on brain injury in DHLF mouse model

Mice with PVT1 overexpression were prepared. qPCR verified the efficiency of PVT1 overexpression in brain tissue. Compared with the control group, lenti-PVT1 significantly increased PVT1 expression (Figure 6a). At 24 h after reperfusion, TTC staining showed that PVT1 overexpression significantly increased the volume of infarcted brain tissue in the DHLF mouse model (Figure 6b, c). At 1 w after reperfusion, MRI showed that PVT1 overexpression significantly increased the volume of infarcted brain tissue (Figure 6d, e) and increased NDS (Figure 6f); meanwhile, open field test suggested that PVT1 overexpression could reduce the total distance of mouse action, decrease the average speed, decreased distance of action in the center, increased distance of action in the periphery, and decreased the number of entries into the center (Figure 6 g-l).

**Figure 6.**
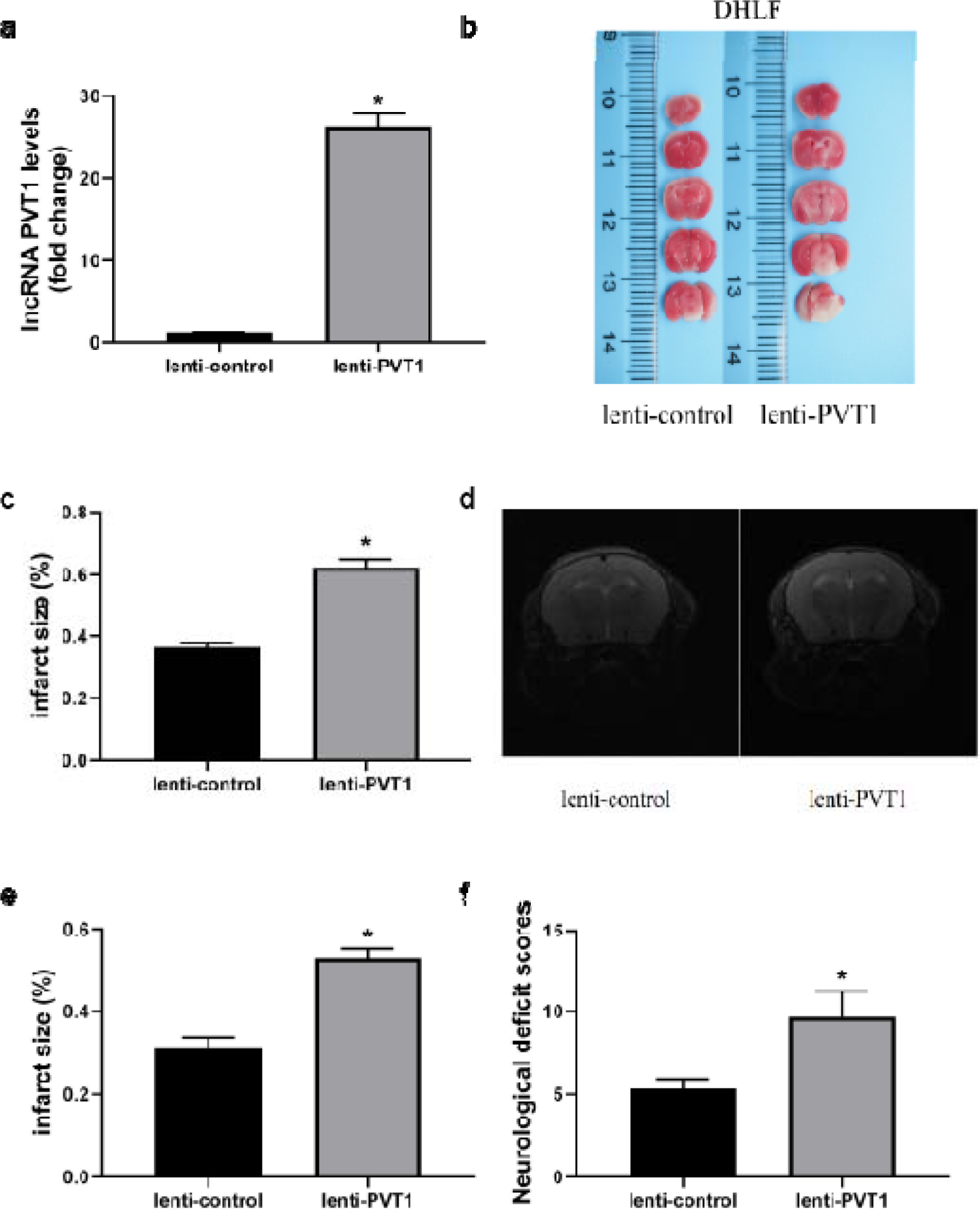
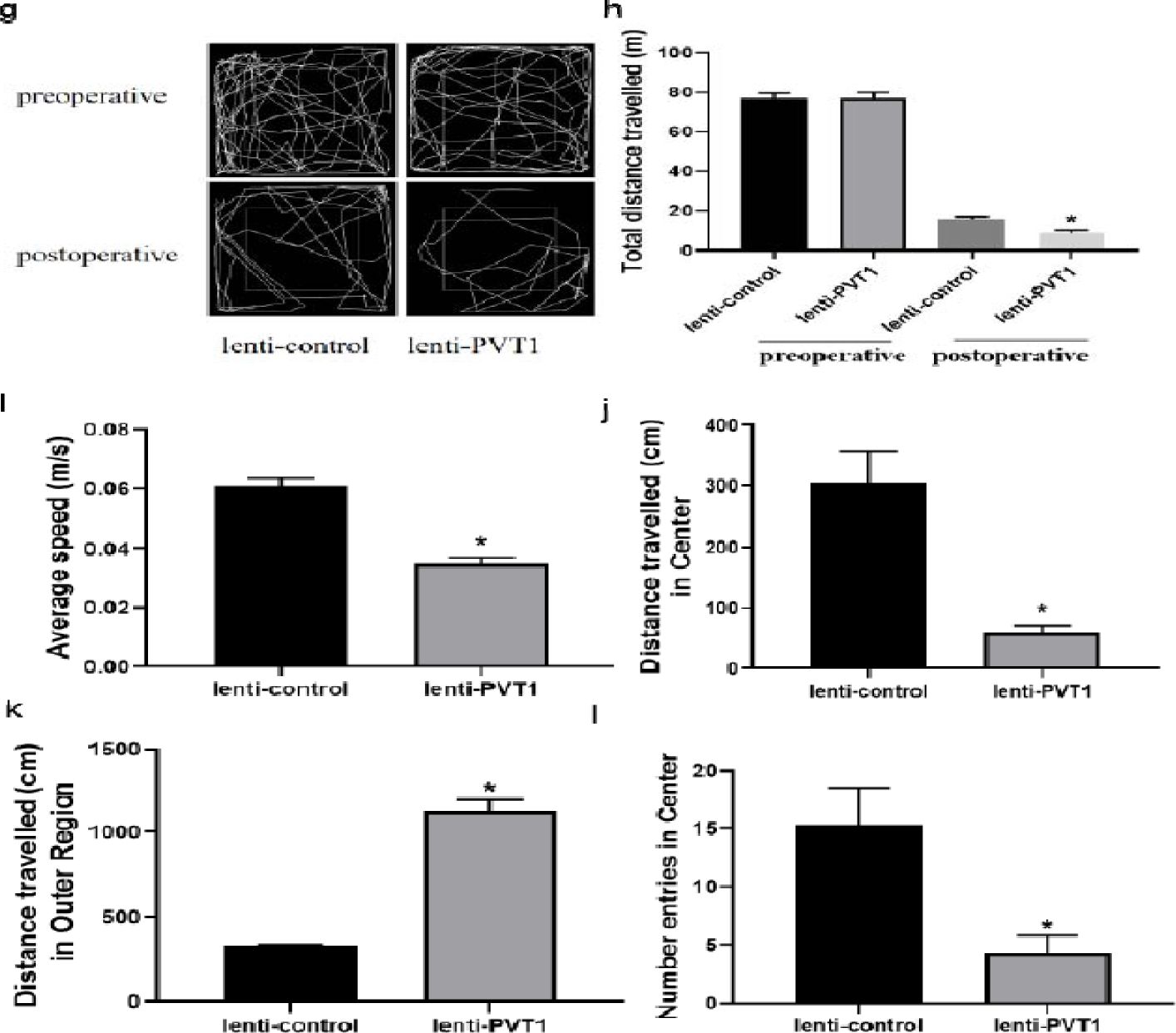
Effect of overexpression of lncRNA PVT1 on brain injury and behavioral changes in DHLF model mice. a: real-time PCR experiments to detect the efficiency of lncRNA PVT1 overexpression in brain tissue (n= 6, *p< 0.05); b&c: TTC staining to assess the volume of brain infarction in mice 24 h after establishment of DHLF model (n= 6, *p< 0.05); d&e: establishment of DHLF model reperfusion 1 week to perform cranial MRI to assess brain damage (n= 6, *p< 0.05); f: establishment of DHLF model reperfusion 1 week to perform deficit score of neurological function (n= 12, * p< 0.05); g-l: establishment of DHLF model reperfusion 1 week to perform absentee field experiment, record the total distance traveled by motor trajectory (g& h), mean velocity (i), distance traveled in the center (j), distance traveled in the periphery (k), and number of times crossing the center (l) (n= 12, * p< 0.05).

### 3.7. Effect of lncRNA-PTV1 overexpression on apoptosis in the deep hypothermia glucose oxygen deprivation cell model

In the PVT1 overexpression hODG-R N2a cell model, the expression of PVT1 was significantly increased in the lenti-PVT1 group compared with that in the lenti-control group (Figure 7a). ELISA results showed that caspase-3 enzyme activity was significantly stronger in the lenti-PVT1 group than in the lenti-control group (Figure 7b). The level of cellular reactive oxygen species (ROS) was significantly higher in the lenti-PVT1 group than in the lenti-control group (Figure 7c). TUNEL assay results showed that the apoptosis rate in the lenti-PVT1 group was significantly higher than that in the lenti-control group (Figure 7d, e).

**Figure 7.**
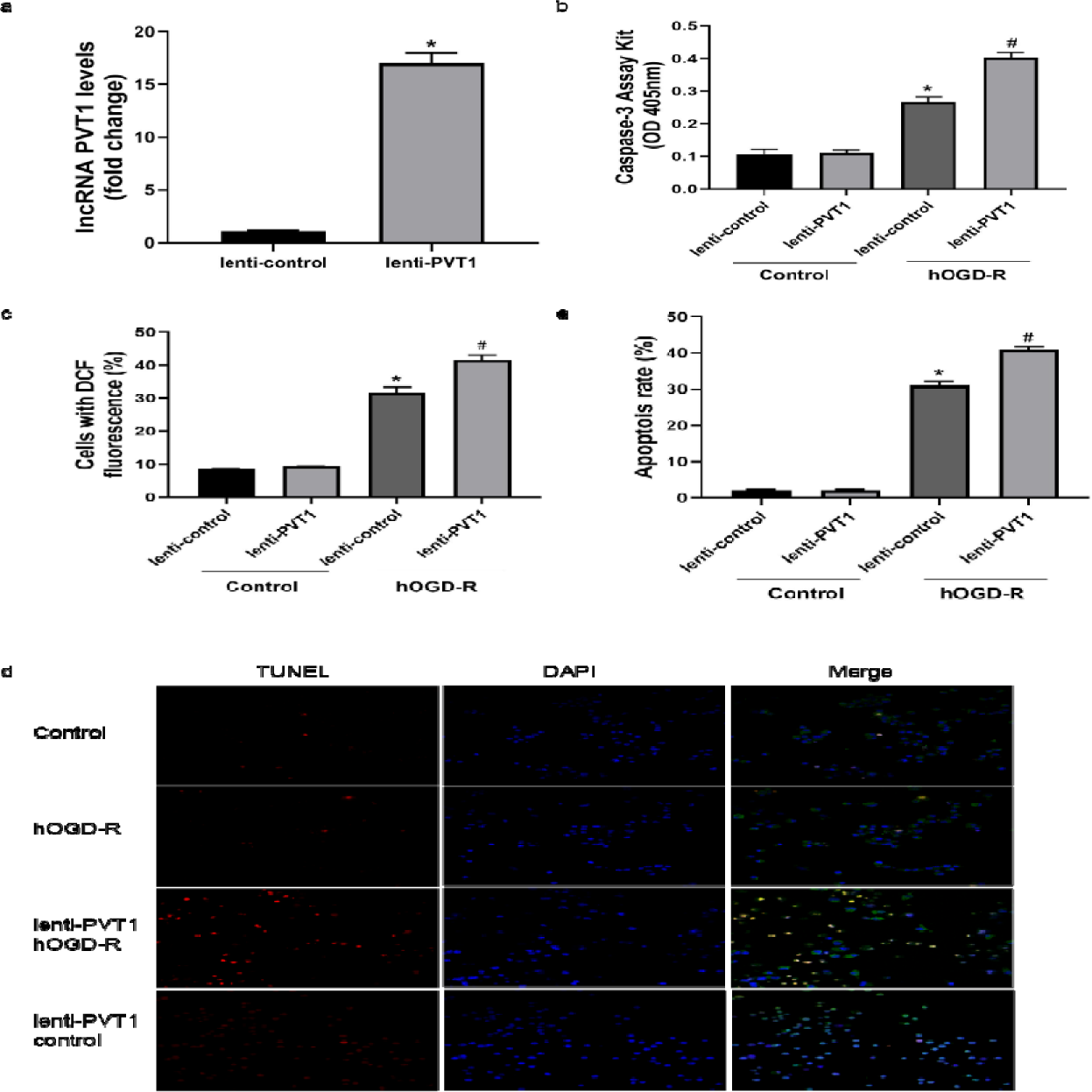
Effect of lncRNA-PTV1 overexpression on apoptosis in a deep hypothermic glucose oxygen deprivation cell model. a: the efficiency of lenti-PVT1 lentivirus vector transfection of N2a cells by real-time PCR (n= 6, *p< 0.05); b: the effect of lenti-PVT1 and lenti control on Caspase-3 activity in N2a cells by caspase-3 activity kit (n= 6, *p< 0.05, vs control group; n= 6, #p< 0.05, vs lncRNA PVT1 overexpression negative control in hOGD-R group); c: DCFH-DA probe to detect ROS production in cells of each group (n= 6, *p< 0.05, vs control group; n= 6, #p< 0.05, vs lncRNA PVT1 overexpression negative control in hOGD-R group); d&e. Immunofluorescence staining showing the effect of lenti-PVT1 and lenti-control lentivirus on apoptosis of N2a cells in control and hOGD-R groups (scale bar: 50 um) and analysis of apoptotic cells in selected regions by Image J software (n= 6, *p< 0.05, vs control group; n= 6, #p< 0.05, vs hOGD-R group in lncRNA PVT1 overexpression negative control group). (control: control group; hOGD-R: a deep hypothermic glucose oxygen deprivation cell model group; lenti-PVT1: lncRNA PVT1 overexpression model; lenti-control: lncRNA PVT1 overexpression model negative control)

### 3.8 Effect of PVT1 overexpression on apoptosis in the DHLF mouse model

To study the role of PVT1 in the DHLF model, a DHLF model with PVT1 overexpression was constructed. TUNEL and immunofluorescence assays showed apoptosis in the DHLF group (Figure 8a, b). When PVT1 was overexpressed, the rate of apoptosis was increased in the lenti-PVT1 group, compared to that in the negative control group (Figure 8 a&b). Further ROS fluorescence staining showed a higher level of ROS in the DHLF group than in the sham group. When PVT1 was overexpressed, the level of ROS was significantly increased in the lenti-PVT1 group, compared to that in the negative control group (Figure 8 c&d). ELISA assay results showed that compared to that in the sham group, Caspase-3 activity was increased in the DHLF group (Figure 8 e). When PVT1 was overexpressed, Caspase-3 activity was significantly increased in the lenti-PVT1 group compared to that in the negative control group (Figure 8 e).

**Figure 8.**
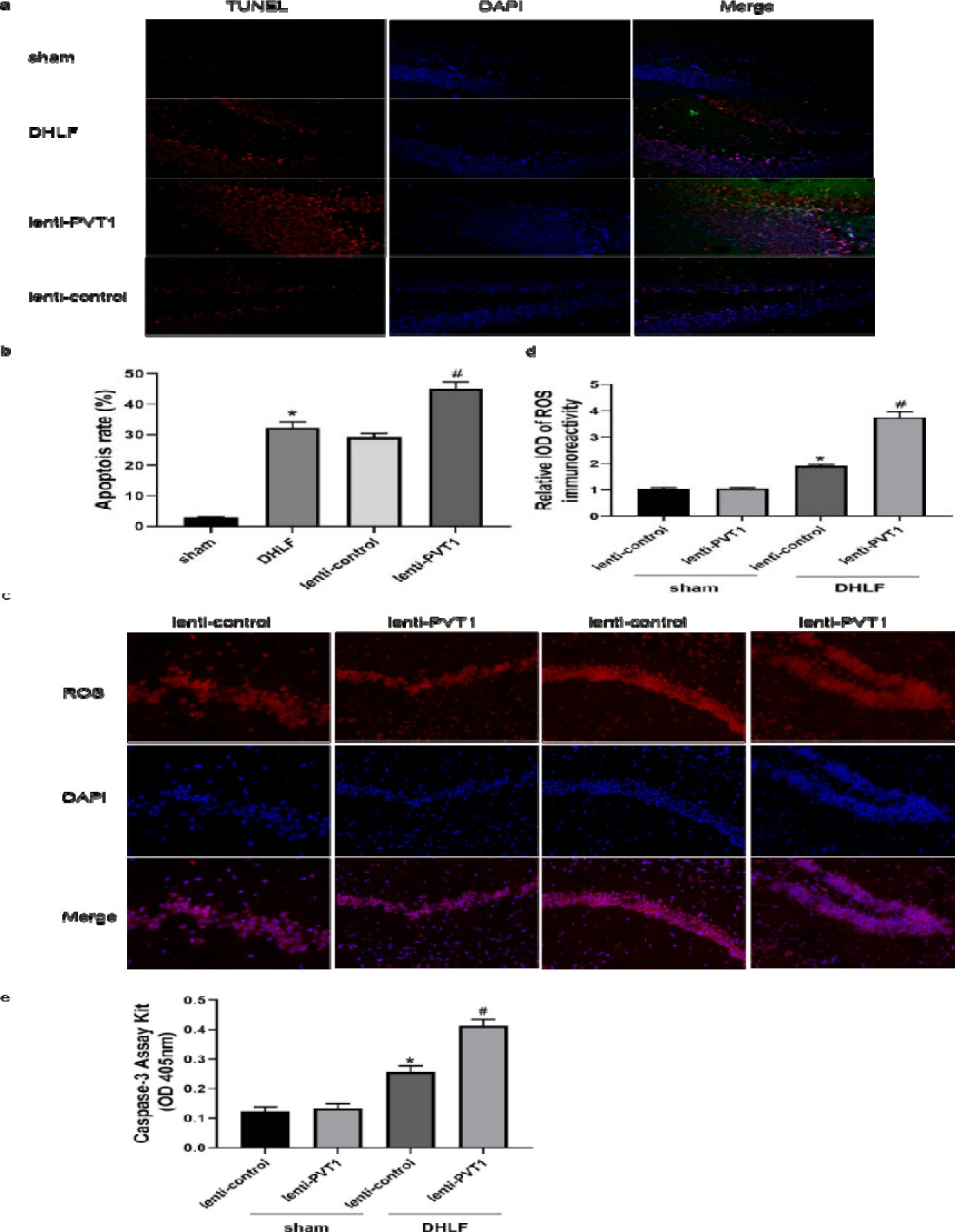
Effect of lncRNA PVT1 overexpression on apoptosis in brain tissue of DHLF mouse model. a&b: Immunofluorescence staining showing the effect of lenti-PVT1 and lenti-control lentivirus on apoptosis in brain tissue of sham and DHLF groups (scale bar: 50 um) and apoptotic cell analysis of selected regions by Image J software (n= 6, *p< 0.05, vs sham group; #p< 0.05, vs lncRNA PVT1 overexpression negative control group); c&d: immunofluorescence staining showing the effect of lenti-PVT1 and lenti-control lentivirus on ROS levels in brain tissues of mice in sham and DHLF groups and by Image J software calculated (n= 6, *p< 0.05, vs sham group; #p< 0.05, vs lncRNA PVT1 overexpression negative control group); e: ELISA to detect the effect of lenti-PVT1 and lenti-control lentivirus on Caspase-3 enzyme activity in brain tissues of mice in sham and DHLF groups (n= 6, *p< 0.05, vs sham group; #p< 0.05, vs lncRNA PVT1 overexpression negative control group). (sham: sham-operated group; DHLF: deep hypothermic low flow group; lenti-PVT1: lncRNA PVT1 overexpression model; lenti-control: lncRNA PVT1 overexpression model negative control).

### 3.9 Interaction between PVT1 and miR-148a-3p

Bioinformatics analysis (RNAhybrid, TargetScan) was performed to predict which miRNA binds to PVT1. Comparing the sequences of PVT1 and miR-148a-3p, we found that the PVT1 sequence contained possible binding sites of miR-148a-3p (Figure 9 a). Meanwhile, the dual luciferase reporter gene assay showed that miR-148a-3p mimic inhibited luciferase activity in HEK293T cells transfected with vector containing wild-type PVT1 reporter gene (Figure 9 b). qPCR results showed that compared with that in the control group, the expression of mir-148a-3p was significantly increased in the hOGD cell model, brain tissue of DHLF mice and blood samples of children undergoing DHLF (Figure 9 c-e).

**Figure 9.**
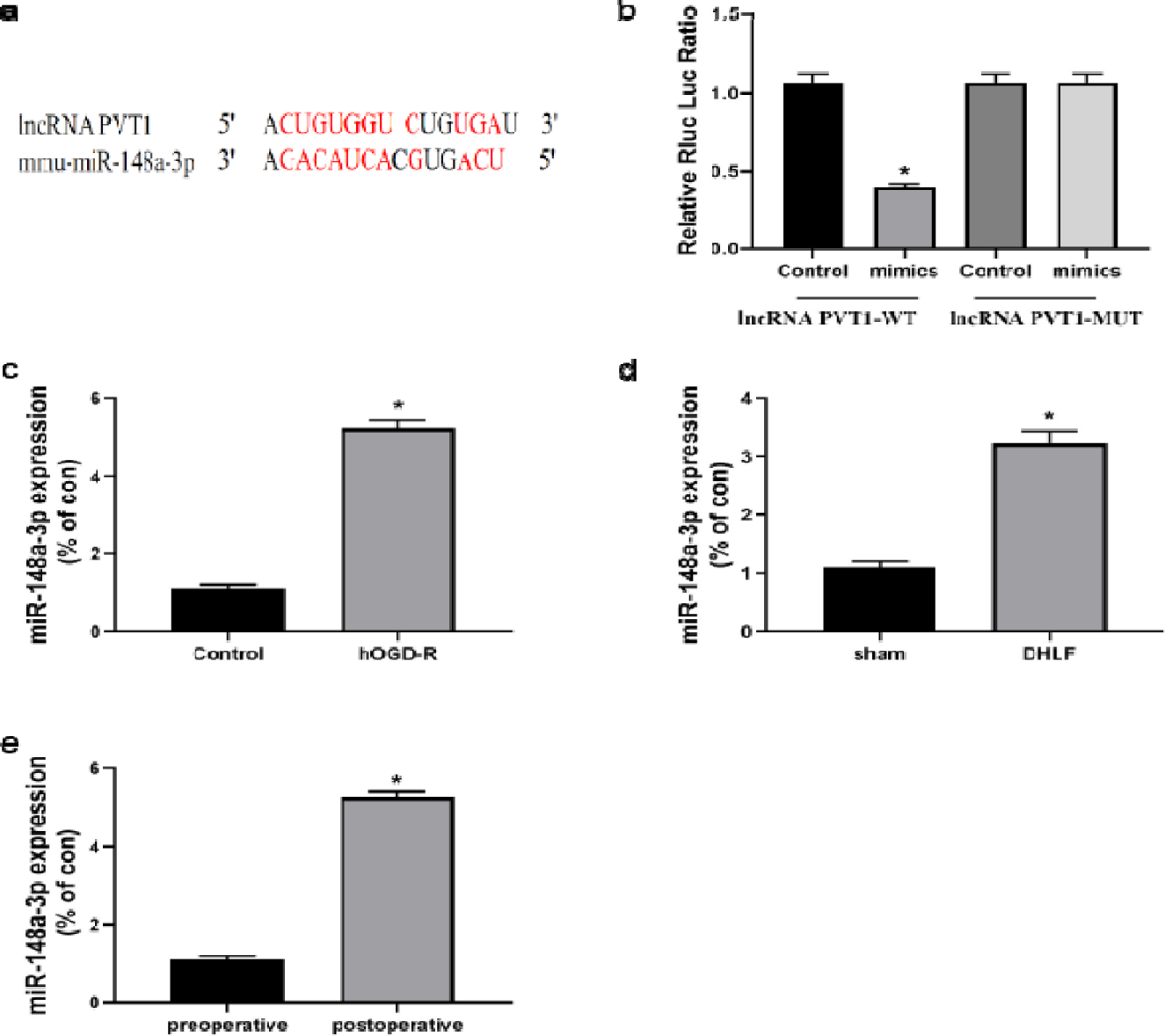
lncRNA PVT1 interacts with miR-148a-3p. a: bioinformatic analysis to predict the binding site of lncRNA PVT1 to miR-148a-3p; b: luciferase reporter gene assay to verify the binding of lncRNA PVT1 to miR-148a-3p (n= 6, p< 0.05); c: real-time PCR to detect the expression of miR-148a-3p in the N2a cell hOGD-R model (n= 6, *p< 0.05); d: real-time PCR to detect the expression of miR-148a-3p in the brain tissue of mouse DHLF model (n= 6, *p< 0.05); e: real-time PCR to detect the in vitro process on miR-148a-3p expression in the blood of the children (n= 6, *p< 0.05). (control: control group; hOGD-R: deep hypothermic glyoxylation deprivation-reoxygenation cell model group; DHLF: mouse deep hypothermic low-flow group; lenti-PVT1-WT: lncRNA PVT1 genotype as wild-type group; lncRNA PVT1-MUT: lncRNA PVT1 genotype as mutant group).

### 3.10 Interaction between miR-148a-3p and MKL1

Analysis with TargetScan and miRwalk software showed that the 3□-UTR of MKL1 sequence might be a binding site of miR-148a-3p (Figure 10 a). Dual luciferase reporter gene assay showed that miR-148a-3p mimic and wild-type MKL1 3□-UTR of pmiR-GLO plasmid co-transfected with HEK293T cells could down-regulate luciferase activity (Figure 10 b). qPCR and Western blotting showed that in the hOGD cell model, brain tissue of DHLF mouse model and blood samples of children undergoing DHLF, the protein and mRNA levels of MKL1 expression decreased (FIG. 10 c-k).

**Figure 10.**
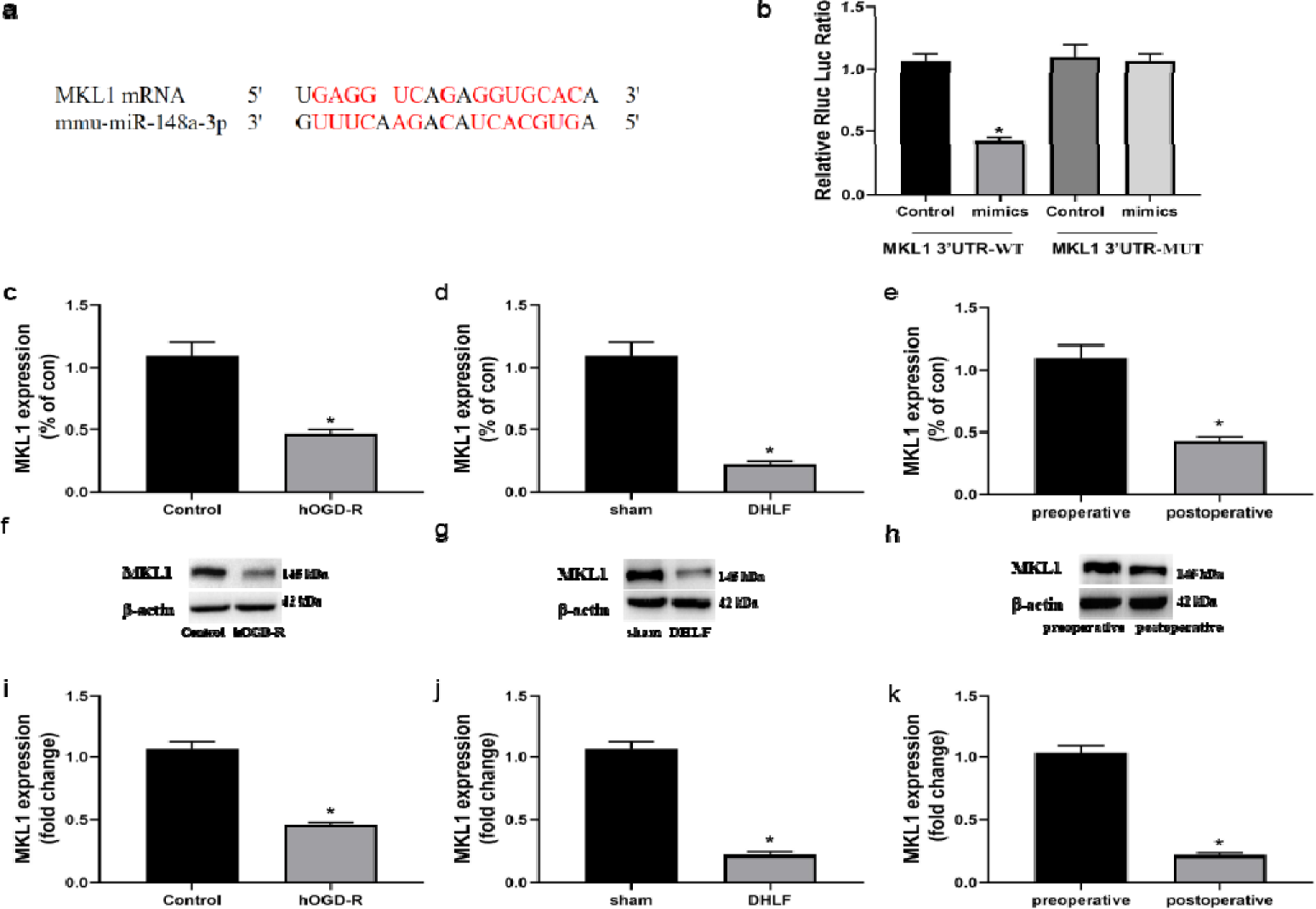
MKL1 interacts with miR-148a-3p. a: bioinformatic analysis to predict the possible binding sites of MKL1 mRNA to miR-148a-3p; b: dual luciferase reporter gene assay to verify the binding of miR-148a-3p to MKL1 mRNA 3’UTR (n= 6, *P< 0.05); c-e: real-time PCR analysis of MKL1 mRNA expression in hOGD-R-treated N2a cell model, mouse DHLF model brain tissue, and blood samples from children undergoing DHLF extracorporeal circulation, respectively (n= 6, *P< 0.05); f-k: western blot analysis of MKL1 protein in hOGD-R-treated N2a cell model, mouse DHLF model brain tissue, and blood samples from children undergoing DHLF extracorporeal circulation (n= 6, *P< 0.05), respectively. (control: control group; hOGD-R: deep hypothermic glyoxylation deprivation-reoxygenation cell model group; MKL1 3’UTR-WT: MKL1 genotype is wild type; MKL1 3’UTR-MUT: MKL1 genotype is mutant).

### 3.11 MKL1 as a downstream target of PVT1/miR-148a-3p axis

N2a cells containing agomir-148a-3p (FIG. 11 a and c) and antigomir-148a-3p (FIG. 11 b and d) were transfected to establish the hOGD model. It was found that mir-148a-3p was overexpressed in the agomir-148a-3p group. Meanwhile, compared with that in the control group, the expression of MKL1 protein decreased, while the expression of mir-148a-3p was lower in antigomir-148a-3p group. In the hOGD cell model (FIG. 11 e & g) and mouse DHLF model (FIG. 11 f & h), the expression level of MKL1 protein in PVT1 overexpression group increased, compared with that in the control group. In order to verify that Mkl1 is a downstream target, a MKL1 function blocking model was constructed (DHLF model was established by intraperitoneal injection of Mkl1 inhibitor CCG1423, 1 mg/kg/d for 1 week). The immunofluorescence staining and TUNEL staining showed that in the two models, compared with the antigomir-148a-3p group, the apoptosis rate decreased in MKL1 inhibitor group (Mkl1 inhibitor group: N2a cells were cultured with CCG1423 24 hours before the establishment of the model for 1 day) (FIG. 11 i & j; m & n). Compared with that in the control group, the apoptosis rate in the lenti-PVT1 group decreased after administration of MKL1 inhibitor (FIG. 11 k & l; o & p).

**Figure 11.**
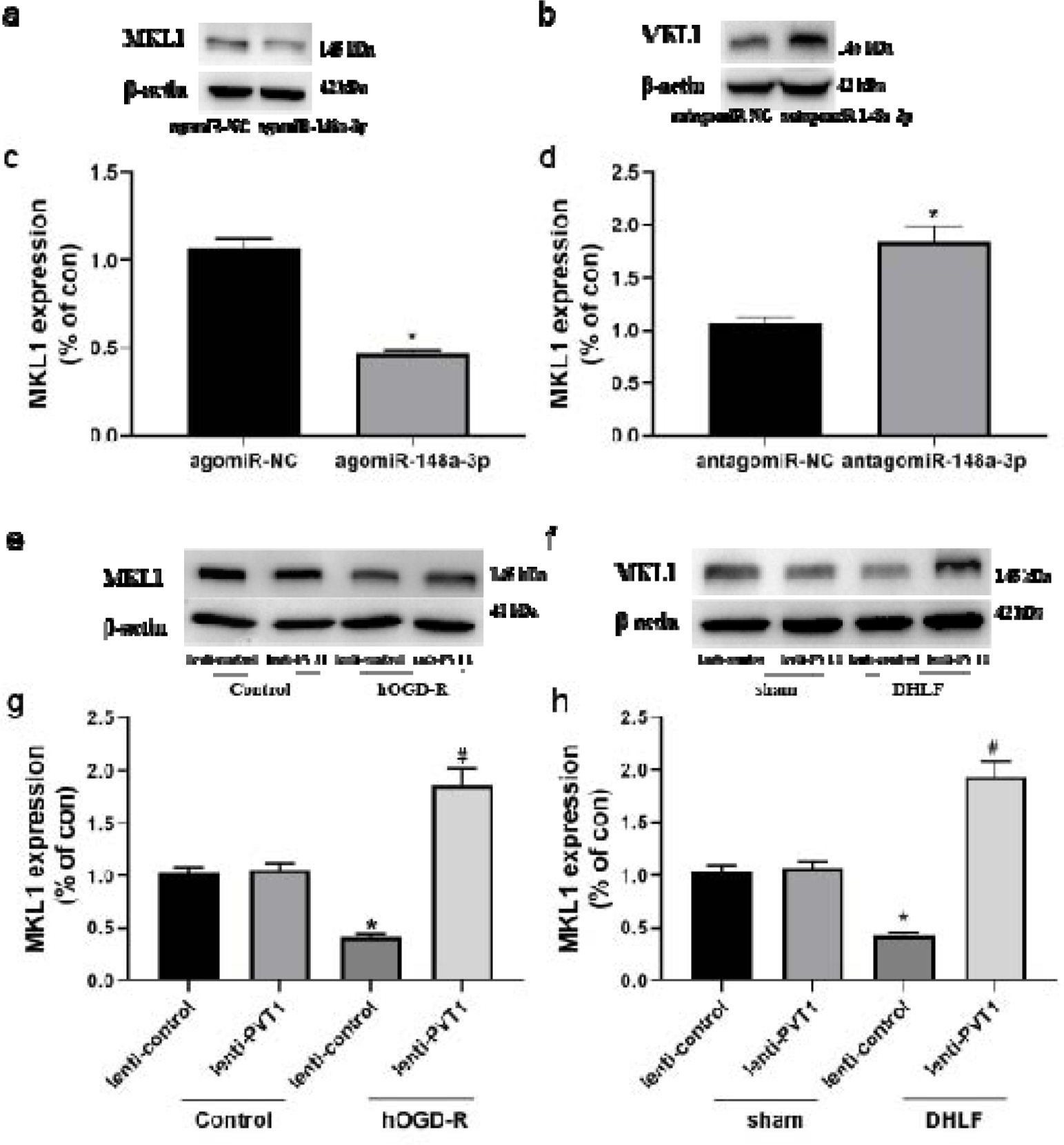
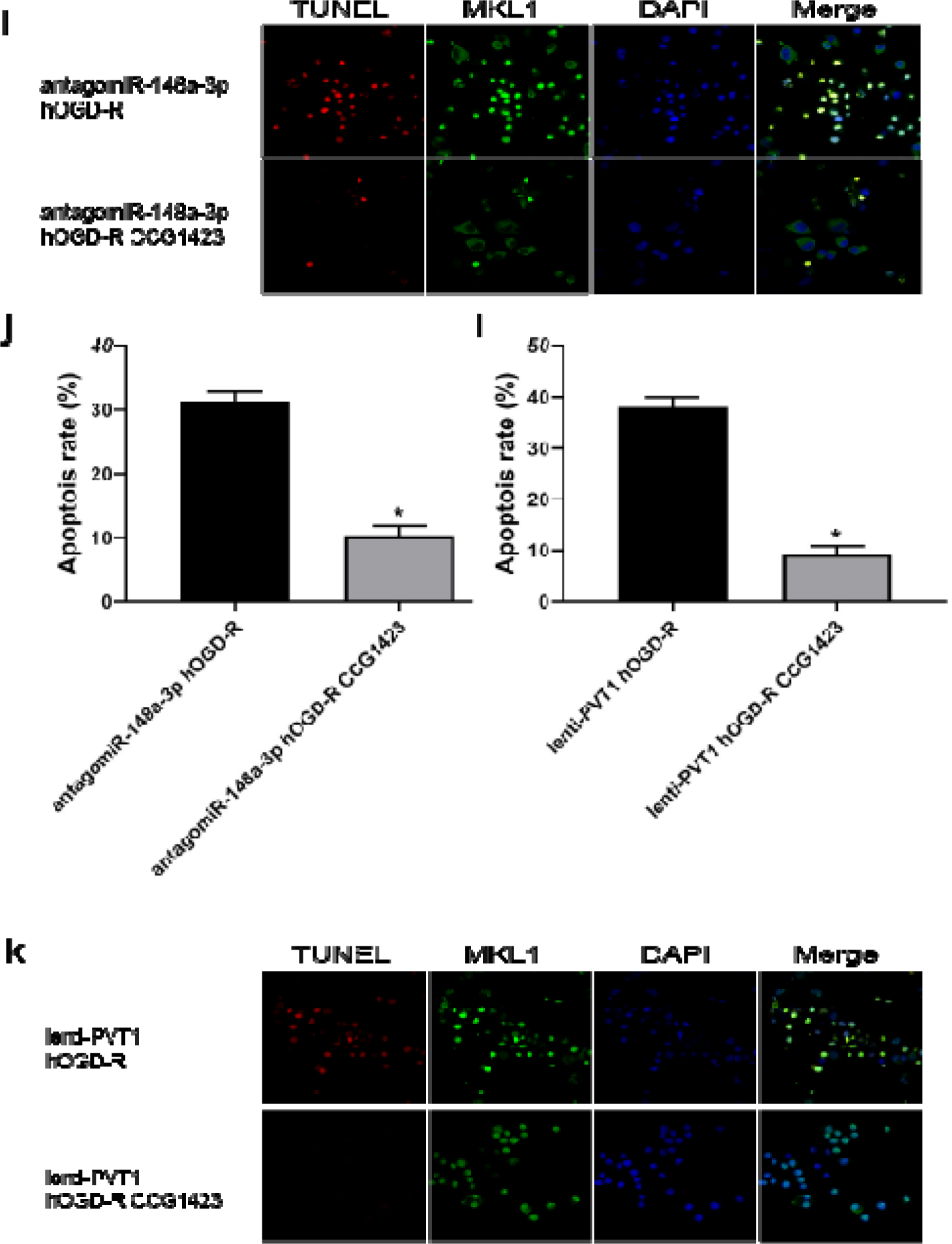
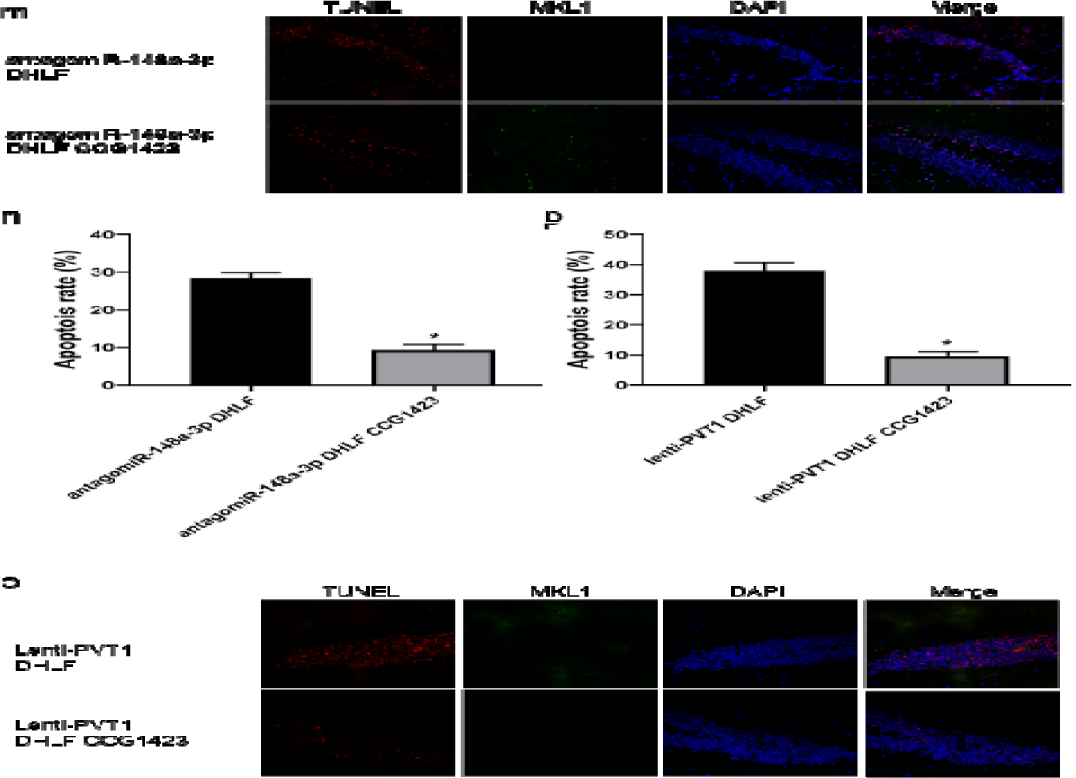
MKL1 as a downstream target molecule of lncRNA PVT1/miR-148a-3p axis. a&c: agomiR-NC, agomiR-148a-3p transfected N2a cells, established hOGD-R model, western blot to detect the protein expression level of MKL1 in cells (n= 6, *p<0.05); b&d. antagomiR-NC, antagomiR-148a-3p transfected N2a cells, hOGD-R model established, western blot to detect the protein expression level of MKL1 in the cells (n= 6, *p<0.05); e&g: western blot to detect the effect of lenti-PVT1 and lenti-control viral vectors on MKL1 protein expression in cells of control and hOGD-R groups (n= 6, *p< 0.05, vs control group; n= 6, #p<0.05, vs hOGD-R model in lncRNA PVT1 overexpression negative control group); f&h: western blot to detect the effect of lenti-PVT1 and lenti-control viral vectors on MKL1 protein expression in brain tissue of mice in sham and DHLF groups (n= 6, *p< 0.05, vs sham group; n= 6, #p<0.05, vs DHLF model in lncRNA PVT1 overexpression negative control group); i&j: immunofluorescence analysis of apoptosis in antagomiR-148a-3p-transfected N2a cells hOGD-R model after MKL1 inhibitor blockade (n= 6, *p<0.05); k&l: immunofluorescence analysis of apoptosis in lncRNA PVT1 after MKL1 inhibitor blockade apoptosis in the overexpression group after MKL1 inhibitor blockade (n=6, *p<0.05); m&n: immunofluorescence analysis of apoptosis in the brain group of antagomiR-148a-3p lateral ventricle injected mouse DHLF model after MKL1 inhibitor blockade (n=6, *p<0.05); o&p: immunofluorescence analysis of apoptosis in the brain group of lenti-PVT1 slow apoptosis in the mouse DHLF model brain group after viral lateral ventricular injection (n=6, *p<0.05). (control: control group; hOGD-R: deep hypothermic glyoxylation deprivation-reoxygenation cell model group; lenti-PVT1: lncRNA PVT1 overexpression model group; lenti-control: lncRNA PVT1 overexpression model negative control group; agomiR-148a-3p: miR-148a-3p overexpression group. agomiR-NC: miR-148a-3p overexpression negative control; antagomiR-148a-3p: miR-148a-3p silent expression group; antagomiR-NC: miR-148a-3p silent expression negative control group).

### 3.12 Neuronal apoptosis regulated by PVT1 through the miR-148a-3p/MKL1 axis

To further verify that PVT1 regulates MKL1 expression via miR-148a-3p, agomiR-148a-3p and PVT1 overexpression lentivirus vectors were co-transfected in N2a cells. The results showed that PVT1 overexpression significantly inhibited the decrease of MKL1 expression caused by agomiR-148a-3p (Figure 12 a&b). Meanwhile, knockdown of PVT1 reduced the increase of MKL1 expression induced by antagomiR-148a-3p (Figure 12 c&d). After transfection of N2a cells with pcDNA3 MKL1 effectively increased the expression of MKL1 protein (Figure 12 e&f). Meanwhile, flow cytometry showed that apoptosis rate was increased in the hOGD-R group, and MKL1 overexpression increased apoptosis rate (Figure 12 g&h).

**Figure 12.**
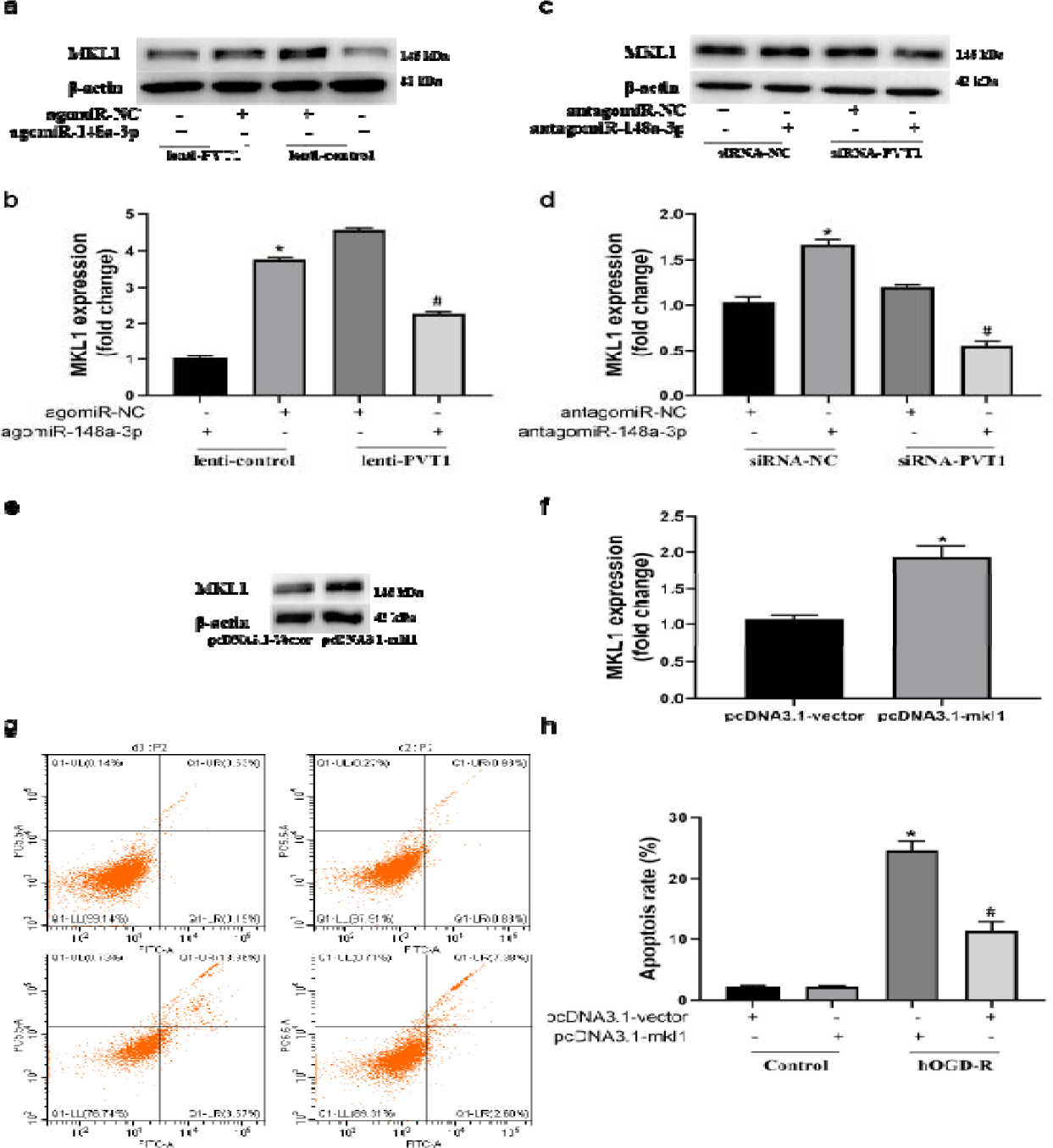
The lncRNA PVT1/miR-148a-3p axis regulates apoptosis through MKL1. a&b: western blot detection of MKL1 protein expression in N2a cells after co-transfection with lenti-PVT1 lentivirus and agomiR-148a-3p (n= 6, *p<0.05, vs agomiR-NC group. n= 6, #p<0.05, vs agomiR-148a-3p group); c&d: western blot detection of MKL1 expression in N2a cells after co-transfection with siRNA-PVT1 and antagomiR-148a-3p (n= 6, *p<0.05, vs antagomiR-NC group; n= 6, #p< 0.05, vs antagomiR-148a-3p group); e&f: Western blot detection of MKL1 protein expression after transfection with pcDNA3.1-MKL1; g&h: flow cytometry detection of the effect of N2a cells transfected with pcDNA3.1-MKL1 on apoptosis in hOGD model cells (n= 6, *p<0.05 vs pcDNA3.1-vector group; n= 6, #p<0.05, vs control group). (control: control group; hOGD: deep hypothermic glyoxylation deprivation/reoxygenation cell model group; lenti-PVT1: lncRNA PVT1 overexpression model; lenti-control: lncRNA PVT1 overexpression model negative control group; agomiR-148a-3p: miR-148a-3p overexpression group. agomiR-NC miR-148a-3p overexpression negative control; antagomiR-148a-3p: miR-148a-3p silent expression group; antagomiR-NC: miR-148a-3p silent expression negative control group; pcDNA3.1-vector: MKL1 overexpression negative control group. (pcDNA3.1-mkl1: MKL1 overexpression group; siRNA-NC: lncRNA PVT1 silent expression negative control group; siRNA-PVT1: lncRNA PVT1 silent expression group).

## 4. Discussion

In the surgery under extracorporeal circulation of CHD children, DHLF is often performed at body temperature of 18°C-20°C and flow rate of 20-25 ml/kg/min to provide essential blood for vital organs [20, 21]. However, despite this protective effect, many organs, especially the central nervous system, may still confront IRI during long-term DHLF. Even after treatment with DHLF, up to 30% of patients have a stroke or other damage to the brain [16, 22, 23]. Therefore, the mechanism of brain IRI after DHLF should be clarified to provide basis for the invention of treatment options.

In recent years, the relationship between IRI and ncRNAs has been increasingly studied. According to their structures, ncRNAs are classified into miRNAs, long-stranded non-coding RNAs (lncRNAs) and circular RNAs (circRNAs) [24], which regulate various biological processes in cells [25]. Among them, lncRNAs consist of more than 200 nucleotides and play critical roles in cell cycle, gene cleavage, mRNA degradation, and translation. Recently, Wu et al. reported that 2292 lncRNAs were upregulated and 1848 downregulated in patients with myocardial infarction [26]. Liu et al. performed gene chip analysis in a mouse model of myocardial I/R, finding that 64 lncRNAs were upregulated and 87 downregulated in the infarcted region [27]. In a cell-based study, 309 lncRNAs were upregulated and 488 downregulated in H9c2 cells following hypoxia/reoxygenation [28]. These findings demonstrate that lncRNAs are involved in IRI. It has reported that lncRNAs act in the pathophysiology of brain IRI [29]. lncRNAs are dysregulated in cerebral ischemia to induce neuronal death, microglia activation, inflammation and neurogenesis. In ischemic stroke, the expression of lncRNA transcripts and the activation of signal transduction pathways are cell-type-specific [30]. After ischemic stimulation, lncRNA MALAT1 expression increases by about 6-fold in both in vitro and in vivo middle cerebral artery occlusion (MCAO) experiments [31,32]. lncRNA MALAT1 knockout mice exhibit a large infarct size, a poor neurological score and severer sensorimotor dysfunction [33]. In the brain tissue upon IRI, the expression of lncRNA H19 is elevated. lncRNA H19 upregulation increases the levels of ROS, malondialdehyde and lactate dehydrogenase, as well as inhibits P13K/AKT phosphorylation [33]. lncRNA MEG3 overexpression enhances the transcriptional activity of p53, leading to apoptosis and necrosis [34]. These show that lncRNAs are closely related to IRI.

Clinical practice has revealed that long-term DHLF may cause severe damage to the CNS damage throgh I/R, and there are no effective prevention and treatment options [34]. In this study, 72 differentially expressed lncRNAs between DHLF and control groups, including 38 up-regulated and 34 down-regulated, with the most significant change in PVT1, which is located at 8q24.21[35]. The expression of PVT1 in the blood samples from children experiencing DHLF and hOGD cells was also consistent with the sequencing results. Aberrant expression of PVT1 has recently been reported in many human cancers. PVT1 can be used as a non-invasive diagnostic biomarker for small cell lung cancer (NSCLC) [36]. PVT1 promotes angiogenesis [37]. PVT1 is downregulated to regulate neuroelectric activity and apoptosis in the hippocampus, a mechanism that may involve autophagy-related pathways [38]. PVT1 is upregulated in the pathogeneses of some cardiovascular diseases [39]. Knockdown of PVT1 inhibits the apoptosis of vascular smooth muscle cells and the destruction of extracellular matrix in abdominal aortic aneurysms [40]. PVT1 protects human AC16 cardiomyocytes from H/R-induced apoptosis [41,42]. Therefore, PVT1 may be downregulated to inhibit the apoptosis of cells and protect from IRI.

In the present study, PVT1 overexpression significantly increased the volume of infarcted brain tissue and the NDS in the DHLF mouse model. Meanwhile, open field test confirmed that PVT1 overexpression could reduce the motor ability of mice. This suggests that PVT1 plays an important role in the development of brain IRI. PVT1 overexpression also appeared with Caspase-3 activity enhancement, suggesting that PVT1 may employ Caspase-3 to regulate apoptosis. The same phenomenon was observed in the hOGD cell model. Therefore, we hypothesize that PVT1 regulates downstream molecules to facilitate apoptosis in brain IRI. Brain injury was exacerbated when PVT1 was overexpressed, implying that inhibition of PVT1 may attenuate brain IRI by reducing apoptosis, which provides a new direction for the prevention and treatment of brain IRI.

It has been reported that in a complex RNA regulatory network, lncRNA exerts a de-repressive effect on target mRNAs by competitively binding to microRNAs [43]. lncRNAs can serve as the sponge of microRNAs [44]. PVT1 can also directly sponge miRNAs to regulate cancer cell development. PVT1 acts as a competitive endogenous RNA for miR-497 and promotes NSCLC progression [45]. Yang et al. found that PVT1 sponged miR-365 to inhibit ATG3 expression in hepatocellular carcinoma[46]. Downregulation of PVT1 inhibited glycolysis and progression of osteosarcoma cells and attenuated the expression of HK2 sponged by miR-497 [47]. Through the VEGFA/VEGFR1/AKT axis, PVT1 sponges miR-16-5p to promote colorectal carcinogenesis [48]. PVT1 promotes ovarian carcinogenesis and metastasis by interacting with miR-140 [49]. These results reveal that PVT1 interacts with miRNAs or protein-coding genes to regulate cancer progression. Serum PVT1 expression increases significantly in patients with acute ischemic stroke [50]. In the present study, we further verified the molecular mechanisms of PVT1 in brain IRI.

Bioinformatics analysis and sequence analysis revealed that PVT1 might bind to miR-148a-3p. In the DHLF mouse model group, miR-148a-3p was lowly expressed but PVT1 was overexpressed when brain tissue damage was aggravated, suggesting a negative association between the expression of PVT1 and miR-148a-3p. High miR-148a-3p expression was associated with alleviation of brain tissue injury. Therefore, it is hypothesized that miR-148a-3p is also involved in the development of brain IRI after DHLF. miR-148a-3p is derived from miR-148a and both share similar activities [51]. miR-148a reduces liver injury in mice by regulating calmodulin-dependent protein kinase II. miR-148a may regulate the MAPK pathway during hypoxia and glucose deficiency-induced microglia inflammation, which is of critical importance in hypoxia-induced ischemic injury. lncRNA-H19 acts endogenously and competitively with miR-148a-3p to inhibit ROCK 2 activity and thus oxidative stress. However, the expression and function of miRNA-148a-3p in the DHLF mouse model have not been reported. To verify the relationship between PVT1 and mir-148a-3p in brain IRI, carrying wild-type 3’UTR lncRNA (LUC lncRNA PVT1-3’utr-wt) and 3’UTR binding site mutation (LUC lncRNA PVT1-3’utr MUT) was designed. Dual luciferase reporter assays showed that co-transfection of miR-148a-3p with Luc-PVT1-3’UTR-WT significantly reduced the relative luciferase activity, and Luc-PVT1-3’UTR-Mut binding site mutation could not affect the luciferase activity. This suggests that PVT1 can target miR-148a-3p. Together, PVT1 may act as a sponge of miR-148a-3p in the process of brain IRI after DHLF.

In the single cell RNA-sequencing analysis of myofibers, PVT1 was found to be a regulator of mitochondrial respiration, fission/fusion and mitochondrial phagocytosis/autophagy [52]. IRI involves mitochondrial dysfunction, calcium overload, leukocyte infiltration and inflammation, and energy depletion, all contributing to the accumulation of ROS during IRI through different mechanisms [53]. We have confirmed in previous experiments [2,3,17] that ROS level was significantly upregulated in the brain tissue of the normothermic mouse IRI model (which differs from the DHLF model in that the body temperature of the mouse was not lowered when the bilateral internal carotid arteries were clamped), compared to the that in the DHLF model group, along with an increase in brain damage. It is evident that oxidative stress is involved in brain IRI after DHLF. In mammals, the NADPH oxidase (NOX) family plays a key role in the production of ROS. In the present study, bioinformatic analysis predicted the possible binding sites of miR-148a-3p to MKL1. In cardiac macrophages, MKL1 recruits the acetyltransferase MOF of histone H4K16, which stimulates NOX gene transactivation and promotes myocardial IRI [54]. However, pharmacological inhibition of the MKL1-MOF-NOX axis (CCG1423 is an inhibitor of MKL1) can attenuate myocardial IRI in mice. MKL1 acts as a trigger of oxidative stress. MKLl is expressed in many types of tissue and usually functions as a coactivator of serum response factors [54]. In this study, a DHLF model was established by intraperitoneal injection of MKL1 inhibitor CCG1423 (1 mg/kg/d for 1 week). In the hOGD cell model, the antagomiR-148a-3p group showed higher MKL1 protein expression and apoptosis rate, and this rate was reversed after administration of MKL1 inhibitor (N2a cells were challenged with CCG1423 culture for 24 h before model building). MKL1 protein expression level and apoptosis rate increased in the PVT1 overexpression group, and this rate was reversed after administration of MKL1 inhibitor. The same results were obtained in the DHLF mouse model. Therefore, PVT1, MKL1 and miR-148a-3p cooperate to regulate apoptosis of neuronal cells.

In order to verify the relationship between miR-148a-3p and MKL1, wild-type 3’UTR MKL1 mRNA (Luc-MKL1-3’UTR-WT) and mutated 3’UTR (Luc-MKL1-3 ’UTR-Mut) were constructed. Dual luciferase reporter assay showed that co-transfection of miR-148a-3p with Luc-MKL1-3’UTR-WT significantly reduced the relative luciferase activity, and Luc-MKL1-3’UTR-Mut could not change luciferase activity. This suggests that miR-148a-3p can bind to MKL1, which is a downstream target of miR-148a-3p. Meanwhile, PVT1 overexpression significantly suppressed the decrease in MKL1 expression caused by agomiR-148a-3p. Meanwhile, knockdown of PVT1 reduced the MKL1 expression upregulated by antagomiR-148a-3p. PVT1 overexpression significantly attenuated the protective effect of agomiR-148a-3p on cells, as shown by the increased apoptosis rate. Together, PVT1 may function by regulating the miR-148a-3p/MKL1 axis in DHLF-induced brain I/R.

## Conclusions

1. A total of 72 lncRNAs with differential expression were identified in brain tissues of DHLF model mice. The expression of PVT1 in the DHLF mouse model, hOGD cell model and blood of children undergoing DHLF showed a similar profile, indicating that PVT1 may be involved in the process of brain injury.
2. PVT1 may serve as an effective molecular therapy to reduce brain injury after DHLF in children treated with surgery under extracorporeal circulation.
3. PVT1 regulates oxidative stress and apoptosis through the miR-148a-3p/MKL1 axis.
4. Inhibition of PVT1 may alleviate brain IRI by repressing apoptosis, which provides a new direction for the prevention and treatment of brain IRI.

## Limitations

1. In this study, DHLF was experimented as an overall condition. They can be studied experimentally as intervention factors separately if necessary.
2. Other lncRNAs regulating the function of the miR-148a-3p/MKL1 axis in a direct or indirect manner should be investigated.
3. Whether MKL1 directly regulates the function of PVT1 under DHLF conditions requires further experimental studies.
4. Whether PVT1 can be a biomarker for the diagnosis, treatment, and prognosis of postoperative brain injury after DHLF requires clinical studies with large sample sizes.
5. The findings should be validated in brain tissue specimens.

## Data Availability

All data mentioned in the manuscript are original.

## Acknowledgements

We want to thank Dr siyu ma and xiaodong zangfor scientific and technical assistance.

Conflict of interest: none declared.

